# Costs associated with increasing numbers of Medicare beneficiaries with HIV aged 65 years and older from 2026 to 2035

**DOI:** 10.64898/2025.12.19.25342703

**Authors:** Emily P. Hyle, Luke Ang, Grace Luu, Parastu Kasaie, Dannie Dai, Satoshi Koiso, Jessica Phelan, Florence Ebem, Ciara Duggan, Elizabeth Humes, Paul E. Sax, Lucas Gerace, John Giardina, E. John Orav, Anne M. Neilan, Ankur Pandya, Jose F. Figueroa, Keri N. Althoff, Kenneth A. Freedberg

## Abstract

**Importance:** As the population of older people with HIV (PWH) in the US is growing, costs to Medicare are expected to rise substantially.

**Objectives:** To project the number of Medicare beneficiaries with HIV aged 65y+ on ART in the US from 2026-2035 and the budget impact on Medicare.

**Design, Setting, and Participants:** We developed the novel CHARMED simulation model and projected the number of Medicare beneficiaries with HIV aged 65y+ on ART and associated costs from 2026 to 2035; we populated the model with age and sex-stratified clinical data and costs derived from 2023 Traditional Medicare (TM) claims and accounted for enrollment in Medicare Advantage, as well as healthcare inflation.

**Main Outcomes and Measures:** Numbers of Medicare beneficiaries with HIV aged 65y+ on ART and undiscounted costs to Medicare from 2026-2035.

**Results:** We projected that 111,600 PWH would be enrolled in Medicare and in care at the beginning of 2026 (65-69y: 57,370; 70-74y: 32,940; 75-79y: 14,670; 80y+: 6,610). By the end of 2035, this number would nearly double, to 193,560 (65-69y: 70,490; 70-74y: 62,820; 75-79y: 38,290; 80y+: 21,960). Annual costs to Medicare for PWH 65y+ on ART would increase 2.5-fold, from $11.4 billion at the end of 2026 to $28.6 billion at the end of 2035. Cumulative 10-year costs are projected to be $195.6 billion with 66.5% of cumulative costs due to ART. If ART costs are reduced by 60% as per the Inflation Reduction Act or generic ART, Medicare would save $78.0 billion over the next decade.

**Conclusions and Relevance:** The number of Medicare beneficiaries with HIV 65y+ on ART will more than double over the next decade, resulting in $195.6 billion in 10-year total costs to Medicare. Reducing ART costs through the IRA or generic oral ART could lead to 40% lower overall Medicare spending for older Medicare beneficiaries with HIV.

**KEY POINTS:** *Question:* As the population of people with HIV in the US grows older, what are the expected costs to Medicare and the impact of antiretroviral therapy (ART) costs?

*Findings:* Using microsimulation modeling, we find that the number of Medicare beneficiaries with HIV 65y+ on ART will more than double over the next decade. At current costs of ART and health care-associated inflation, total 10-year costs to Medicare are anticipated to be $196.5 billion; reducing ART costs through the Inflation Reduction Act or generic ART could lower Medicare spending by 40%.

*Meaning:* Efforts to reduce ART costs, while maintaining access to high-quality ART, are critical to reduce total Medicare costs as more people with HIV are anticipated to enroll in Medicare in the next 10 years.

## INTRODUCTION

With contemporary antiretroviral therapy (ART), the life expectancy of people with HIV (PWH) in the United States has improved markedly and is now near normal.^1–3^ Most PWH aged 65 years or older are enrolled in Medicare, either qualifying due to eligible conditions prior to age 65y or through age-eligibility at 65y.^4^ This aging population is expected to grow substantially over the next decade, ^5,6^ and medical complexity and clinical care costs will rise with a high burden incurred by Medicare. However, the extent of that increase remains uncertain.

Moreover, rising ART costs and their overall contribution to Medicare costs for PWH are unknown.^7^ The Inflation Reduction Act (IRA), passed in 2022, offers a new opportunity for the federal government to negotiate and lower the price of drugs for Medicare.^8^ One of the proposed drugs for price negotiation, bictegravir/emtricitabine/tenofovir alafenamide (Biktarvy), is an ART regimen that is estimated to have amassed over $3.1 billion in Medicare spending during 2023 alone.^9^ Additionally, a generic first-line ART regimen that is highly effective is expected to become available in 2031 (e.g., dolutegravir and lamivudine).^10^ Reducing ART costs among PWH in Medicare offers an important opportunity to decrease the lifetime costs of the growing population of PWH in Medicare without compromising clinical care or outcomes. Our goal was to project the numbers of PWH aged 65 years or older on Medicare in the US over the next decade and the costs of their care.

## METHODS

### Analytic Overview

Using the Cost-Effectiveness of Preventing AIDS Complications (CEPAC-US) model, a previously validated microsimulation model of HIV disease, we projected the total numbers and age distributions of PWH in the US from 2026-2035, assuming current trends in the care continuum.^11–14^ We developed the Cardiovascular, HIV, Aging, heaRing loss, MEntal health, and Dementia (CHARMED) model, a novel multimorbidity simulation model of clinical and economic outcomes among PWH aged 65y+ who are engaged in clinical care in the US. We populated the CHARMED model with CEPAC projections of the age distribution and population size of PWH 65y+ at the beginning of January 2026, as well as the number of PWH turning aged 65y each month from 2026-2035. Using the CHARMED model, we limited the simulated population to Medicare beneficiaries aged 65y+ with HIV on ART and then projected the annual population size and age distribution from 2026 to 2035, as well as annual total costs incurred over 10 years by Medicare, accounting for inflation. We performed sensitivity analyses to examine the uncertainty of parameter estimates and their effects on model outcomes.

This analysis was approved by the Mass General Brigham human research committee, who waived the need for informed consent, as no individual-level data were used in the models. We used the Consolidated Health Economic Evaluation Reporting Standards (CHEERS) 2022 reporting guidelines (eTable 1 in **Supplement 1**).^15^

### The CEPAC Model

We populated the CEPAC-US model for adults 20y+ with CDC surveillance data from 2014 (eTable 2 in **Supplement 1**). We then calibrated projected transmissions to CDC HIV incidence data from 2014-2022, reflecting the impact of rising PrEP use from 2014-2022 (eMethods and eTable 3 in **Supplement 1**). ^16–19^ We compared CEPAC model projections to CDC surveillance data for 4 distinct outcomes: numbers of people living with diagnosed HIV, percent of people receiving care and with virologic suppression, and deaths. ^20–23^

### The CHARMED Model

The CHARMED Model is a multimorbidity microsimulation model that simulates clinical care for people aged 65y+ on ART and projects clinical and cost outcomes. At model start, simulated individuals are in care and on ART as per national guidelines; we assume that people remain in care throughout their lifetime.^24^ Each individual draws from pre-specified distributions for clinical characteristics, as well as a likelihood of attaining virologic suppression on ART and experiencing virologic non-suppression due to resistance or intermittent adherence. People diagnosed with virologic non-suppression can re-suppress with re-initiation of ART or a change to a new ART regimen. Each month, people are at risk for death due to: HIV-related mortality (e.g., cryptococcal meningitis) and non-HIV-related mortality (e.g., stroke), which is age-/sex-stratified and reflects the increased risks of non-communicable diseases among PWH.^25^

### CHARMED Model Cohorts

To project the numbers of PWH aged 65y+ on ART enrolled in either Traditional Medicare or Medicare Advantage from 2026-2035, we constructed three cohorts in the CHARMED model: Medicare beneficiaries aged 65y+ in care in 2026; PWH turning age 65y each month after 2026 who are Medicare-enrolled and in care; and Medicare-enrolled people aged 65y+ initiating ART after a new HIV diagnosis. We weighted these three cohorts to create the population of all Medicare beneficiaries with HIV aged 65y+ on ART and then projected their clinical outcomes and costs (eMethods in **Supplement 1**).

### CHARMED Model Inputs

Using CMS enrollment and CDC data, we estimated that 74% of PWH 65y+ in care are enrolled in Traditional Medicare with Part D or Medicare Advantage coverage (eMethods in **Supplement 1**);^16^ the other 26% are either not given employer-sponsored insurance or are unable to acquire and maintain Medicare coverage (e.g., permanent residency requirements, prohibitive costs, work credit requirements). ^26–28^ Taking the CEPAC-projected number of PWH 65y+ at the end of 2025, we estimated 112,050 Medicare beneficiaries with HIV at CHARMED model start (e.g., beginning of 2026), in four age categories: 57,710 PWH aged 65-69y, 32,930 (70-74y), 14,700 (75-79y), and 6,710 (80y+) (**Table 1**). The Medicare-enrolled population in care is 77% male with a mean (SD) CD4 count of 576/µL (266/µL) for PWH in care, and 363/µL (203/µL) for people newly diagnosed with HIV and initiating care.^29,30^ Throughout the simulation, Medicare beneficiaries with HIV enter the cohort monthly: Medicare-enrolled PWH who are newly 65y+ (694 to 925 people/month) and newly diagnosed PWH entering care aged 65y+ (37 people/month) **(Table 1)**.^31^ We estimated that 90% of PWH in care will attain virologic suppression,^23^ and we incorporated both HIV-related and non-HIV-related mortality.^12,25,32^

**Table 1.** Selected base case input parameters for Medicare beneficiaries aged 65y+ on ART in the CHARMED Model.

| Parameter | Base Case Input | Reference |
| --- | --- | --- |
| Baseline cohort characteristics |  |  |
| Male at birth (%) | 77 | 60 |
| Initial CD4 count, mean cells/μL (SD) | | |
| PWH in care age 65+ | 576 (267) | 61 |
| PWH in care turning 65y | 576 (267) | 61 |
| Newly diagnosed with HIV age 65+ | 363 (203) | 62 |
| Total number of PWH in care in January 2026 |  |  |
| 65-69 years | 50,170 |  |
| 70-74 years | 27,020 | CEPAC-model<br>projections |
| 75-79 years | 11,240 |  |
| $\geq 80$ years | 4,850 | |
| Number of PWH in care turning 65y, monthly | 1,140-2,590 |  |
| Number age 65y+ newly diagnosed with HIV, monthly | 70 | 31 |
| PWH 65y+ engaged in care who are Medicare-enrolled (%) | 82 | 16 |
| PWH 65+ engaged in care who are virologically suppressed (%) | 90 | 23 |

**Table 1. Selected base case input parameters for Medicare beneficiaries aged 65y+ on ART in the CHARMED Model. (CONT.)**
| Parameter | Base Case Input |  | Reference |
| --- | --- | --- | --- |
| Costs |  |  |  |
| Mean annual spending per person (\$) | | | 34 |
| Male |  |  |  |
| Age | Each 12 months of survival | 12 months prior to death |  |
| 65-69 years | 81,491 | 165,638 |  |
| 70-74 years | 79,633 | 159,767 |  |
| 75-79 years | 80,686 | 161,856 |  |
| ≥ 80 years | 83,392 | 140,486 |  |
| Female |  |  |  |
| 65-69 years | 81,494 | 170,126 |  |
| 70-74 years | 78,455 | 149,400 |  |
| 75-79 years | 78,776 | 132,343 |  |
| ≥ 80 years | 79,079 | 156,308 |  |

Abbreviation: PWH; People with HIV, CHARMED: HIV, Aging, heaRing loss, MEntal health, and Dementia microsimulation model

**Table 2.** Model-projected base case age-stratified numbers of Medicare beneficiaries aged 65y+ alive at year end and annual age-stratified costs.

| Number of Medicare beneficiaries aged 65y+ alive at year end |  |  |  |  |  | Costs in billions of USD* |  |  |  |
| --- | --- | --- | --- | --- | --- | --- | --- | --- | --- |
| Year | 65-69y | 70-74y | 75-79y | 80y+ | All 65y+ | ART | Direct<br>HIV-related | Other<br>Non-HIV-related | Total<br>costs |
| 2025 | 57,370 | 32,940 | 14,670 | 6,610 | 111,600 | - | - | - | - |
| 2026 | 60,390 | 36,340 | 17,200 | 7,970 | 121,890 | 7.2 | 0.3 | 4.3 | 11.4 |
| 2027 | 63,190 | 39,870 | 19,650 | 9,230 | 131,930 | 8.2 | 0.4 | 4.8 | 13.0 |
| 2028 | 66,400 | 43,240 | 21,910 | 10,280 | 141,830 | 9.4 | 0.4 | 5.3 | 14.6 |
| 2029 | 69,610 | 46,460 | 23,930 | 11,120 | 151,120 | 10.7 | 0.5 | 5.8 | 16.4 |
| 2030 | 73,200 | 49,380 | 25,590 | 11,680 | 159,850 | 12.0 | 0.5 | 6.3 | 18.3 |
| 2031 | 73,400 | 52,020 | 28,380 | 14,470 | 168,270 | 13.4 | 0.5 | 6.9 | 20.2 |
| 2032 | 73,540 | 54,450 | 31,180 | 17,030 | 176,200 | 14.9 | 0.6 | 7.5 | 22.2 |
| 2033 | 73,420 | 57,200 | 33,790 | 19,130 | 183,540 | 16.5 | 0.6 | 8.0 | 24.4 |
| 2034 | 73,380 | 59,900 | 36,220 | 20,800 | 190,300 | 18.1 | 0.7 | 8.5 | 26.5 |
| 2035 | 70,490 | 62,820 | 38,290 | 21,960 | 193,560 | 19.7 | 0.7 | 9.0 | 28.6 |
| 2026-2035 | - | - | - | - | - | 130.0 | 5.3 | 60.2 | 195.6 |

\*Note: Results are rounded. Values across sub-categories and years may not add up to total costs due to rounding.

### Costs

To estimate annual age- and sex-stratified clinical care costs for beneficiaries enrolled in Traditional Medicare, we used 2023 Traditional Medicare claims data for PWH enrolled in Part D and who filled at least 10 months of ART prescriptions. We separately estimated costs in each year of life and in the 12 months prior to death, given that clinical care costs are usually substantially higher in the year prior to death (eTables 4 and 5 in **Supplement 1**). The amount paid by Medicare for people enrolled in Medicare Advantage is uncertain but is estimated to be 22% more than their Traditional Medicare counterparts.^33^ We therefore weighted clinical care costs by the proportion of beneficiaries with an HIV diagnosis or any ART prescriptions filled who were enrolled in Traditional Medicare (38.5%) or Medicare Advantage (61.5%) based on January 2022 enrollment (Table 1).^34^ To account for increasing health care costs over time, we applied a 6% annual increase to ART costs based on trends in the annual cost of ART,^7,9^ and increased clinical care costs as per Center for Medicare and Medicaid Service’s projections of the personal health index (2026-2035) (eTable 6 in **Supplement 1**).^35,36^ Undiscounted costs are in 2023 USD.

### CHARMED Model Validation

We validated the CHARMED model projections to 2021-2023 CDC data regarding the number of PWH aged 65y+ living with diagnosed HIV, in care, and virologically suppressed (eMethods in **Supplement 1**).^23,37^

### Sensitivity Analyses

We performed one-way sensitivity analyses to evaluate the impact of parameter uncertainty on model outcomes, varying each parameter estimate individually within a plausible range while holding all other parameters at baseline values (eTable 7 in **Supplement 1)**. We then examined the impact of simultaneously varying parameters in multi-way sensitivity analyses, including combinations of the most influential parameters ascertained from one-way sensitivity analyses. We examined the effects of parameter variation on outcomes of the number of Medicare beneficiaries on ART aged 65y+ and the costs incurred by this population; projected costs will be sensitive to both the total number of projected Medicare beneficiaries on ART and cost inputs. We also performed scenario analyses in which costs are not adjusted for Medicare Advantage or inflation.

## RESULTS

### Base Case

We projected that the population of Medicare beneficiaries with HIV on ART aged 65y+ over the next decade would nearly double, from 111,600 in 2026 to 193,560 in 2035, with increases at every age category over the 10 years: 57,370 to 70,490 (65-69y); 32,940 to 62,820 (70-74y); 14,670 to 38,290 (75-79y), and 6,610 to 21,960 (80 years and older) **(Figure 1A).** The magnitude of rising numbers of Medicare beneficiaries with HIV in each age category further increases at older ages; by 2035, the number of Medicare beneficiaries with HIV on ART ages 65-69y will have increased 1.3-fold, compared with 1.9-fold (70-74y), 2.6-fold (75-79y), and 3.3-fold (80y+). The total number of deaths among Medicare beneficiaries with HIV aged 65y+ would also nearly double, from 5,500 in 2026 to 9,640 in 2035 (eTable 8 in **Supplement 1**). Total healthcare spending by Medicare would increase from $11.4 billion in 2026 to $28.6 billion in 2035, including increases in ART costs ($7.2 billion in 2026 to $19.7 billion in 2035), direct HIV-related care ($0.3 billion in 2026 to $0.7 billion in 2035), and other non-HIV-related costs ($3.9 billion in 2026 to $8.2 billion in 2035) **(Figure 1B)**. Total projected costs for Medicare beneficiaries with HIV 65y+ would be $195.6 billion over the next 10 years.

**Figure 1.**
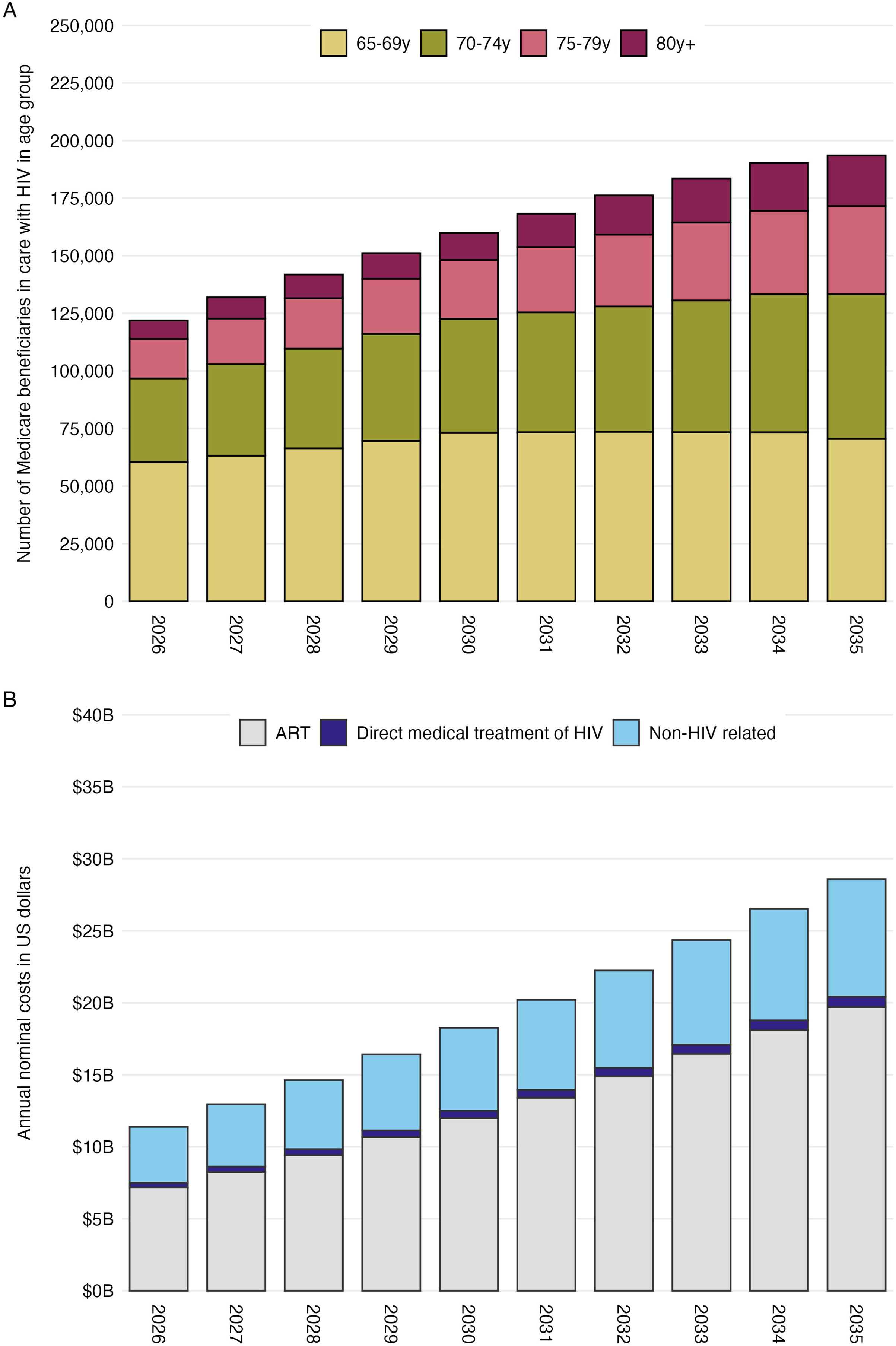
Model-projected numbers and costs of Medicare beneficiaries with HIV aged 65y+ on ART (2026-2035). The age-stratified number of Medicare beneficiaries with HIV on ART by age (Panel A) and the costs to Medicare for this population (Panel B) are projected over the next decade. Panel A displays the number of beneficiaries between the ages of 65-69y (yellow), 70-74y (green), 75-79y (pink), and 80y and older (purple). In Panel B, light blue represents other non-HIV-related costs, dark blue represents costs related to direct medical treatment of HIV, and grey represents ART costs. Abbreviations: ART: antiretroviral therapy.

### Sensitivity Analyses

The most influential parameter in projecting the number of Medicare beneficiaries living with HIV by 2035 is the percentage of PWH aged 65y+ enrolled in Medicare **(Figure 2A)**. If only 68% of PWH aged 65y+ on ART were enrolled in Medicare, there would be only 177,860 Medicare beneficiaries with HIV on ART as of 2035; however, if 94% were enrolled in Medicare, the number of Medicare beneficiaries with HIV on ART would be 245,870 by 2035. Other parameters, including the number of Medicare-enrolled PWH who turn 65y annually and age-/sex-stratified non-HIV-related mortality rates, would have a much smaller impact and would result in 180,000–206,000 Medicare beneficiaries with HIV in 2035.

**Figure 2.**
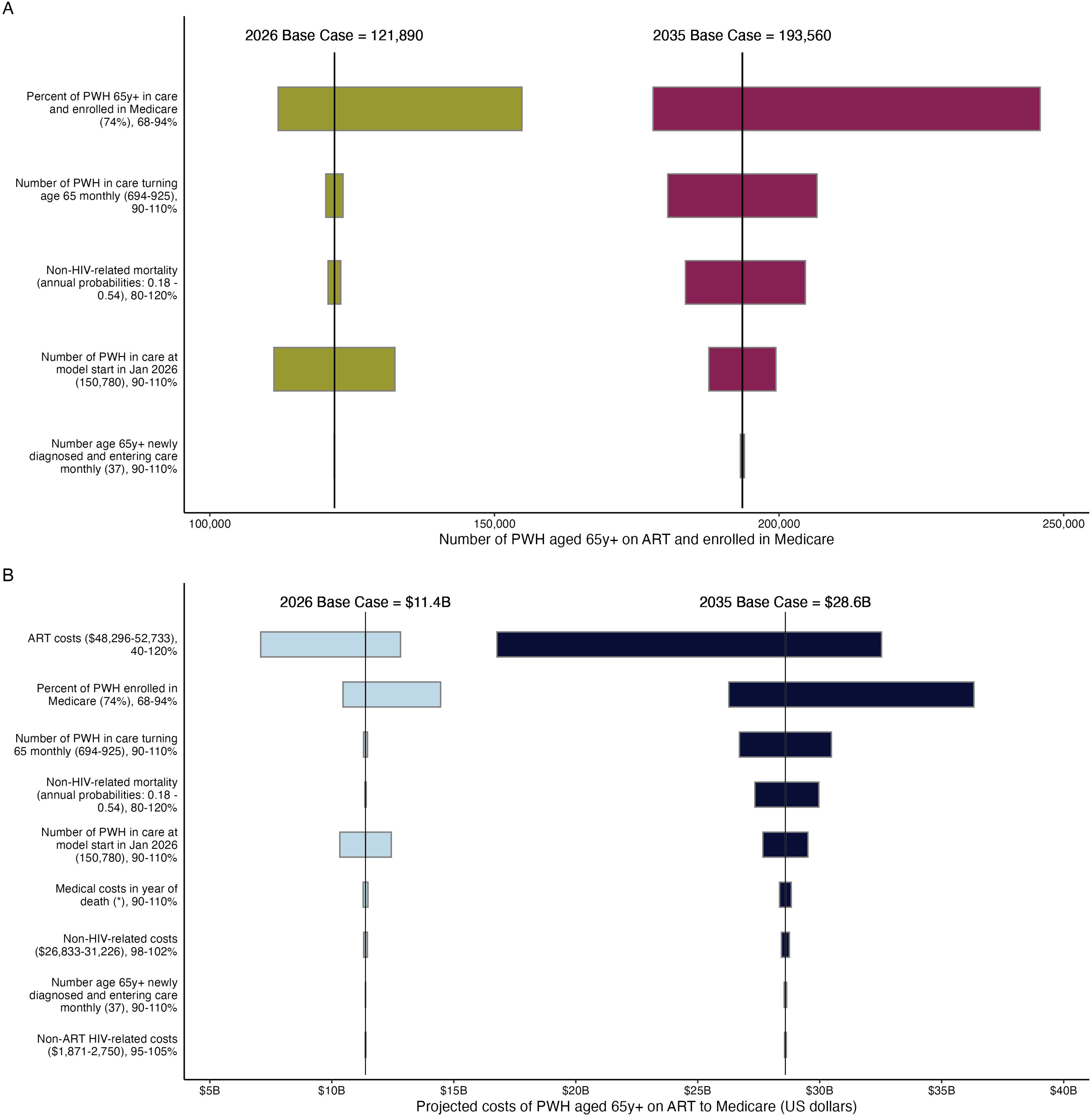
One-way sensitivity analyses on the projected numbers and costs of Medicare beneficiaries aged 65y+ on ART in 2026 and 2035. The age-stratified number of Medicare beneficiaries on ART age 65+ (Panel A) and the costs to Medicare of this population (Panel B) are projected for 2026 (left) and 2035 (right). Input parameters are displayed along the y-axis in descending order from most to least influential. Each parameter is represented by a horizontal bar intersected by a vertical line representing the base case value. The upper bound forms the side of the horizontal bar to the right of the vertical line, and the lower bound forms the side to the left of the vertical line. The one exception in non-HIV-related mortality; when non-HIV-related mortality is higher than the base case, it results in higher costs in 2026 but lower costs in 2035 due to fewer Medicare beneficiaries with HIV aged 65y+ being alive in 2035. Medical costs in the year of death vary depending on cost category, age and sex at birth (eTable 3-4 in **Supplement 1**). Abbreviations: PWH: people with HIV, ART: antiretroviral therapy.

When projecting costs, the most influential parameters are ART costs and the percentage of Medicare-enrolled PWH aged 65y+ on ART **(Figure 2B)**. Reducing per-person ART costs to 40% of 2023 ART costs, as in accordance with the highest statutory minimum discount proposed by the IRA and the estimated impact of generic drug entry (eTable 7 in **Supplement 1**),^38,39^ would reduce Medicare spending to $16.8 billion in 2035 with 10-year cumulative costs of $117.5 billion, whereas an increase of 20% in ART costs would result in $32.5 billion in 2035 with 10-year cumulative costs rising to $221.6 billion (eFigure 1 in **Supplement 1**). Costs to Medicare would range from $26.3-36.3 billion in 2035 if 68-94% of PWH aged 65y+ on ART are enrolled in Medicare. A range in the numbers of Medicare beneficiaries turning 65 years annually, non-HIV-related mortality, and number of Medicare beneficiaries 65y+ in care at model start were less influential on costs in 2035.

In scenario analysis, we also examined the impact of adjusting for increased costs paid for Medicare Advantage patients or health care-associated inflation (eTable 9 in **Supplement 1**). Including a 22% increase in costs of Medicare Advantage beneficiaries results in an increase in 10-year total costs of $23.3 billion. Incorporating health care-associated inflation results in a $62 billion increase in total costs over 10 years, highlighting the impact of anticipated inflation on total costs on Medicare.

In multi-way sensitivity analyses, we examined the impact of different population parameters and non-HIV-related mortality and examined the projected number of PWH Medicare beneficiaries 65+ on ART in 2035 **(Figures 3A and 3B)**. Varying the percentage of PWH 65y+ enrolled in Medicare and non-HIV-related mortality has the most substantial impact: If 94% of PWH aged 65y+ are enrolled in Medicare and non-HIV-related mortality decreases to 80% of current estimates, the number of projected Medicare beneficiaries with HIV would increase to 259,930 in 2035. However, if only 68% of PWH aged 65y+ on ART are enrolled in Medicare and non-HIV-related mortality is 20% higher than current estimates, then only 178,590 PWH aged 65y+ would be enrolled in Medicare in 2035 **(Figure 3A)**. When the number of Medicare-enrolled PWH turning age 65 annually is varied by 10% **(Figure 3B)** with a range in estimated non-HIV-related mortality of 20%, the projected number of Medicare beneficiaries on ART in 2035 would vary between 191,140 and 218,110.

**Figure 3.**
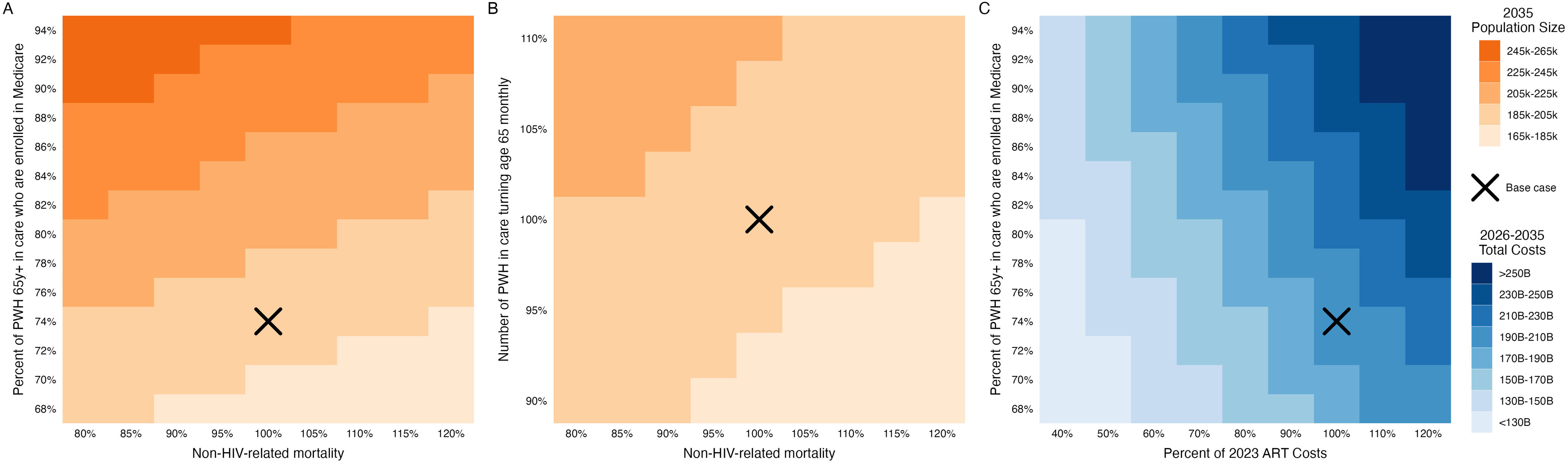
Multi-way sensitivity analyses for the projected population size of and 10-year cumulative costs of Medicare beneficiaries aged 65y+ on ART in 2035. Panels A and B show the model-projected number of Medicare beneficiaries with HIV who are on ART and 65y+ in 2035 given a range in non-HIV-related mortality rates with: 1) percent of PWH age 65y+ who are Medicare-enrolled (Panel A); 2) the number of Medicare-enrolled PWH turning age 65y monthly (Panel B). Panel C shows the model-projected 10-year cumulative costs (2026-2035) given a percent of 2023 ART costs and percent of PWH 65y+ enrolled for Medicare. Light orange (blue) represents a smaller population size (lower costs), whereas darker orange (blue) represents a larger population (higher costs). Base case results are designated with an “X.” Abbreviations: PWH: people with HIV; ART: antiretroviral therapy.

We also varied cost and population parameters to understand the impact of parameter uncertainty on 10-year cumulative costs to Medicare for this population **(Figure 3C)**. If 94% of PWH 65+ on ART are enrolled in Medicare and ART costs increase to 120% of 2023 estimates, then 10-year cumulative costs would be $281.4 billion, in contrast to $108.0 billion if only 68% of PWH 65+ on ART are Medicare-enrolled, and ART costs decrease to 40% of 2023 estimates. Additional multi-way sensitivity analyses are displayed in **eFigure 1 and 2** in the supplementary materials.

## DISCUSSION

In this modeling analysis, we projected that the number of Medicare beneficiaries with HIV aged 65 and older in the United States and engaged in care over the next decade will more than double, reaching almost 200,000 by 2035. The financial burden on Medicare will increase substantially, with annual costs for PWH aged 65y+ increasing from $11.4 billion in 2026 to $28.6 billion in 2035, for a cumulative projected cost of $195.6 billion over 10 years, with ART accounting for approximately 67% of the projected total costs. A 60% reduction in ART costs, such as through drug price negotiations in the Inflation Reduction Act (IRA) or a switch to generic regimens,^38,39^ would result in savings of $78.0 billion over 10 years compared to base case projections.

The growing number of PWH who are Medicare-enrolled based on age is anticipated to affect Medicare substantially, with twice the number of PWH aged 65 years or older insured by Medicare by 2035 compared with 2026. These model-based projections range from 168,670 – 259,930 with plausible changes in the percent of PWH who are Medicare-enrolled at 65 years, non-HIV-related mortality rates, and the numbers of Medicare-enrolled PWH turning 65 years old. Additionally, there is a greater increase in the numbers of Medicare beneficiaries with HIV who reach their 70’s and 80’s, with a 3.3-fold increase in numbers of people older than 80 by 2035 compared with 2026. Improvements in virologic suppression and a reduction in ART toxicities over the past decade support the likelihood that the population of PWH will continue to survive to older ages.^23,40^ Clinicians will need to enhance their understanding of the distinct healthcare needs of this group,^41^ especially given that PWH face an increased risk of comorbidities and are likely to benefit from considerations of multi-morbidity, polypharmacy, drug-drug interactions, and stigma.^42^ The changing healthcare needs of aging PWH will require healthcare systems to adapt to offer comprehensive and person-centered care to tackle the rising burden of comorbidities and healthcare utilization.^43–45^

This model-based analysis underscores that cumulative Medicare costs for PWH could reach nearly $195.6 billion over the next 10 years. With increasing numbers of people living with HIV who are 65 years or older, as well as rising percentage of PWH 65y+ enrolled in Medicare as per RWHAP,^16^ the anticipated increased costs to Medicare over the next decade must be addressed without decreasing the quality or effectiveness of clinical care. Given annual per-person ART costs to Medicare of $47,330-$53,670, ART costs represent the majority of overall spending for Medicare beneficiaries with HIV, exceeding annual costs associated with non-HIV-related clinical care except in the 12 months prior to death. It is, therefore, critical to reduce ART costs to Medicare given that lifetime ART is recommended for all PWH due to individual health benefits and to prevent transmission to others,^24^ and Medicare beneficiaries are not eligible for many rebates and copay options available to people commercially-insured or on Medicaid.^46^ Fortunately, there are now opportunities for ART cost reductions, including the IRA, and an all-generic ART regimen in 2031, when generic dolutegravir becomes available.^47^

Although this analysis provides details on how reduced ART costs would lead to savings to Medicare, policymakers should also consider the interactions between the IRA or generic ART pricing and the 340B program.^48,49^ The 340B program allows for essential care providers for PWH, including disproportionate share hospitals, non-profit organizations, and specific organizations receiving federal funding (e.g., federally qualified health centers and Ryan White clinics), to purchase pharmaceutical drugs at significantly discounted prices and retain revenue that is used for the institution to provide care for underserved populations.^46,49–51^ Current estimates suggest that 340B revenue to participating providers from ART may be decreased by 45-50% through reduced reimbursement rates introduced by the IRA;^48^ similar reductions would likely occur with a switch to prescribing of generic ART. Although there is mixed evidence in the literature as to the impact of the 340B program in increasing access to HIV care,^52–56^ this would lead to a substantial decrease in reimbursement. Further efforts to investigate the impact of reducing ART costs on 340B-enrolled health facilities and subsequent consequences for access to care and health outcomes for PWH will be important. This is particularly relevant since Ryan White Health Care providers and Health Center providers rely heavily on 340B revenues.^46,49^

This analysis has several limitations. First, we limited the scope of model projections to Medicare beneficiaries aged 65y+ to eliminate the heterogeneity introduced by younger PWH enrolled in Medicare given their wide range in health conditions; total costs to Medicare will be even higher when considering Medicare beneficiaries with HIV younger than 65y as noted in KFF estimates.^57^ Mortality among people with HIV at older ages and in care is uncertain and could be lower than our estimates, which would result in a greater number of Medicare beneficiaries with HIV over the next decade, as examined in sensitivity analysis. We used Traditional Medicare data for spending by Medicare and accounted for likely higher costs associated with Medicare Advantage plans,^58^ in which a capitated payment by Medicare is made to private insurers for which the costs to the insurer for clinical care are not public.^59^ To capture the effect of ART costs, we limited the analysis to Medicare beneficiaries with Part D who were engaged in clinical care and on ART; costs to Medicare of people with HIV who are frequently disengaged from HIV care could be higher due to frequent ED visits or lower because of lower ART costs. Because of the focus on Medicare, additional costs of care to Medicaid, commercial insurance, or to the uninsured are not included in this analysis, which includes long-term care costs and cost-sharing borne by Medicaid; however, costs to Medicare of dual-eligible PWH are included in full.

## CONCLUSION

The number of Medicare beneficiaries with HIV older than age 65 in the US will nearly double over the next decade, with almost 193,560 older people insured by Medicare by 2035, resulting in $195.6 billion in cumulative costs to Medicare over the next 10 years. Reductions in ART costs, based on recent legislation and near-term generic availability, could lead to substantial decreases in overall Medicare spending for older beneficiaries with HIV.

## Data Availability

No data were collected for this study. Individual-level CMS data cannot be shared as part of the CMS data use agreement under Dr. Jose Figueroa. However, these data are available for research use through ResDAC if one wishes to pursue their own data use agreement. We are happy to make the input parameterization, including aggregate-level Medicare estimates, and model-based analyses available for review.

