## Supplement 1 for "Costs associated with increasing numbers of Medicare beneficiaries with HIV aged 65 years and older from 2026 to 2035"

**Contents**

**eMethods**

**eResults**

**eReferences**

**eTable 1.** CHEERS 2022 Checklist

**eTable 2.** Input parameters for the simulated cohorts of PWH age 20y+ in the United States in the CEPAC-US Model.

**eTable 3.** Calibration of model transmissions to CDC surveillance estimates.

**eTable 4.** Mean annual per-person costs by category while alive and in the year of death for male Medicare beneficiaries with HIV on ART for 10-12 months of the year.

**eTable 5.** Mean annual per-person costs by category while alive and in the year of death for female Medicare beneficiaries with HIV on ART for 10-12 months of the year.

**eTable 6.** Personal health care index projections used to adjust HIV-related non-ART and non-HIV-related care costs.

**eTable 7.** Parameters assessed in sensitivity analysis: Base case values, lower-bound (LB), upper-bound (UB) and justifications for ranges**.**

**eTable 8**. Validation of CEPAC model output of PWH living with HIV and deaths among PWH diagnosed with HIV aged 20y+ from 2017-2022.

**eTable 9.** Validation of CEPAC model output of receipt of care among diagnosed PWH 20y+ and virologic suppression among PWH 20y+ in care in 2022.

**eTable 10.** Validation of CHARMED model output for the number of PWH over the age of 65, diagnosed, and in care compared with CDC data (2021-2023).

**eTable 11.** Model-projected numbers of deaths among Medicare beneficiaries with HIV aged 65y+ by year-end.

**eTable 12.** Comparison of model-projected costs (in billions of US dollars) based on 4 different approaches to adjusting for higher costs among Medicare Advantage beneficiaries (vs Traditional Medicare beneficiaries) and/or for inflation (vs not).

**eFigure 1.** Impact of changes in ART costs on 10 years of Medicare spending for beneficiaries aged 65y+ on ART (2026-2035).

**eFigure 2.** Multi-way sensitivity analysis of non-HIV-related mortality and the number of Medicare beneficiaries on ART in 2026.

**eFigure 3.** Additional multi-way sensitivity analyses comparing the total cumulative projected costs of Medicare beneficiaries aged 65y+ on ART between 2026 and 2035.

### Supplemental eMethods

#### Using the CEPAC-US model to project incident HIV infections in the US

*CEPAC-US Model Structure*

CEPAC-US is a previously validated microsimulation model of HIV disease, treatment, and transmission.[1] Simulated PWH draw characteristics from distributions of age, sex at birth, HIV RNA, initial CD4 count, and age-stratified adherence to ART at model start.[2] PWH with undiagnosed HIV link to care after HIV testing or opportunistic infection. After linkage to care, PWH initiate ART and can become virologically suppressed, which results in a rise in CD4 cell count, decreased HIV RNA, and decreased HIV-related mortality. Mortality is informed by risk-related behavior among PWH and includes HIV-related causes and non-HIV-related causes. Full specification of the model is provided at https://www.massgeneral.org/medicine/mpec/research/cpac-model.[3]

*Transmission Cohort Structure*

We defined four cohorts of PWH aged 20 and older to reflect the HIV care continuum in the CEPAC model: people newly infected with HIV regardless of diagnosis ("incident" cohort), people living with HIV who are undiagnosed ("undiagnosed" cohort), people diagnosed with HIV receiving ART ("on ART" cohort), and people diagnosed with HIV who are out of care ("disengaged from care" cohort). We used three cohorts (i.e., the undiagnosed, on ART, and disengaged from care cohorts) to project the number and age distribution of people newly infected with HIV ("incident" cohort) based on community viral load,[2] which we then calibrated to CDC data (see Transmission Calibration, eMethods p. 4).

*Transmission Cohort Inputs*

The population is 77% male with a mean (±SD) initial age of 45.8 (12.1) years and a mean (±SD) initial CD4 count of: 400 (166) cells/μL for the undiagnosed cohort, 576 (227) cells/μL for the on ART cohort, 325 (53) cells/μL for the disengaged from care cohort, and 652 (101) cells/μL for the incident cohort (**eTable 2**).[4] We weighted cohorts by the 2014 HIV care continuum as reported by the CDC: PWH who are undiagnosed (151,954), people diagnosed with HIV receiving ART (675,439), and people diagnosed with HIV who are disengaged from care (244,777) (**eTable 2**).[5–7] Additional detailed CEPAC input parameters are included in **eTable 2** and prior publications.[2]

*Transmission Calibration*

To address the trend of decreasing HIV transmissions over the past decade, we applied deflators from 2015 through 2035 to reflect the impact of improving community viral suppression and increasing PrEP use: 1.00 (2015), 0.95 (2016), 0.90 (2017), 0.85 (2018-2019), 0.80 (2020), 0.75 (2021-2022) (**eTable 3**).[8,9]

*CEPAC Model Validation*

We compared model-projected numbers of transmissions among PWH 20y+,[10] the number of people living with diagnosed HIV, and the number of deaths among people with diagnosed HIV from 2017-2022 to CDC reports.[4,10–12] We also compared model-projected percentages of diagnosed PWH 20y+ who receive care and are virologically suppressed at the end of 2022 to CDC estimates.[10]

***Estimating the number of PWH turning 65 annually***

We used the CEPAC model to estimate the number of people living with diagnosed HIV who are 65 years or older in 2026 and who will be turning 65y annually between 2026-2035.

We then estimated the number of PWH aged 65 years or older who are newly diagnosed with HIV using CDC data of new diagnoses of HIV infection among adults aged 65 and older in 2016. In 2016, 844 new diagnoses of HIV infection among people 65y+ were reported, suggesting that a mean of 70 diagnoses are made monthly.[13]

**Validation of the CHARMED Model**

We derived an estimated number of PWH 65y+, diagnosed and engaged in care from CDC estimates for 2021-2023 as a validation target. Then, we compared these derived estimates to the model-projected number of PWH 65y+ who are  diagnosed and engaged in care.[10]

***Projecting the number of PWH age 65y+ enrolled in Medicare***

In this analysis, we projected costs and outcomes only for PWH over the age of 65 who are enrolled in Medicare Part D (and by definition, either Part A and/or Part B) or Medicare Advantage (Part C). Not all PWH aged 65 or older are enrolled in Medicare. Some individuals over the age of 65 are supported with employer-sponsored insurance; others are unable to acquire and maintain Medicare coverage (e.g., permanent residency requirements, prohibitive costs, work credit requirements).[14–16] For some individuals who do not qualify for premium-free Part A due to permanent residency or work credit requirements, Marketplace coverage may be more affordable if their income is between 100-400% of the federal poverty level.[17] In addition, although programs exist to alleviate the Part A, B, and D Medicare premium burden for low-income beneficiaries, regardless of their premium-free eligibility,[18,19] these programs have historically had low participation rates due to high levels of administrative burden and sharp eligibility cutoffs.[20] Individuals who cannot afford the Medicare premiums may opt out (in whole or in part) of Medicare enrollment.[21]

To estimate the percentage of PWH age 65 and older in clinical care who are also enrolled in Medicare, we used a 100% sample of TM and MA beneficiaries in January 2022. We define being in clinical care as having one inpatient or two non-inpatient claims with an HIV diagnosis/DRG code and having any ART filled between 2021-2022 (excluding PrEP). This resulted in 83,484 total PWH 65y+ who were in care, enrolled in Medicare, and taking ART in 2021-2022.

We then derived the number of PWH 65y+ who are diagnosed and in care from the 2022 CDC surveillance and monitoring selected indicator reports.[12,12] We took the proportion of people 65y+ and diagnosed by year-end 2021 who received 1 VL or CD4 test in 2022 (70.7%) and multiplied it by the reported number of PWH 65y+ diagnosed in 2022 from the surveillance report (132,045) to estimate the number of PWH 65y+ who were diagnosed and in care: 113,242 PWH 65y+. Last, we compared the number of PWH 65y+ reported in Medicare (83,484) and divided by the number of PWH 65y+ reported by the CDC to be in care (113,242) and found that 74% of PWH 65y+ in care were also enrolled in Medicare. We assumed that this proportion remains constant between 2026 and 2035.

To derive a range of plausible values for the percent of PWH 65y+ enrolled in Medicare, we used 2022 Ryan White data on client characteristics to estimate the percentage of PWH 65y+ in clinical care who are also enrolled in Medicare.[22] Some Ryan White clients are reported as having “multiple coverages.” Since these likely include both clients with and without Medicare coverage, we included these clients in the calculation of the percentage of clients enrolled in Medicare to get an upper-bound estimate (74%) and also excluded them (61%). We used the midpoint of this range, 68%, as the lower-bound in sensitivity analyses. For the upper-bound of the sensitivity analyses, we used the percentage of the general population over the age of 65y+ who had any form of Medicare coverage in 2022 (94%).[23]

**Parameterization of Medicare Costs**

Using the Medicare dataset of PWH as previously described (see "*Projecting the number of PWH age 65y+ enrolled in Medicare")*, we estimated the age- and sex-stratified clinical care costs for Medicare beneficiaries who were alive for the entire calendar year, excluding any beneficiaries who died in the following 12 months. We separately estimated the clinical care costs of Medicare beneficiaries who died over the 12 months before their death **(eTables 4-5).** To account for the increase in ART costs over time, we applied a 6% annual increase to ART costs based on trends in the annual cost of ART.[24,25] For HIV-related and non-HIV-related clinical care costs, we increased costs as per Center for Medicare and Medicaid Service's projections of the personal health index (2026-2035), with 2023 as a base year **(eTable 6).**[26,27] Undiscounted costs are in 2023 USD.

**Supplementary eResults**

*CEPAC Model Validation*

We projected that 31,870 transmissions would occur in 2022, compared with the calibration target of 31,800 from CDC data **(eTable 3)**.[28]

In 2022, the projected number of diagnosed PWH age 20 or older would be 1,086,840 compared to CDC estimates of 1,087,870. The projected number of deaths among PWH with diagnosed HIV age 20 or older would be 20,730 in 2022 compared with CDC estimates of 18,930 **(eTable 8)**.

We also compare selected outcomes for diagnosed PWH age 20 or older which were available for the year 2022. Receipt of HIV medical care would be 70.6%, as compared to 71.0% per CDC estimates.[29] Viral suppression was projected to be 67.2% compared to 67.2% **(eTable 9)**.[10]

*CHARMED Model Validation*

We projected the total number of PWH aged 65y+ diagnosed and in care from 2021-2023 and validated our projections to estimates derived from CDC data. For 2023, we projected that 128,419 PWH 65y+ would be diagnosed and in care, as compared to 125,419 as derived from CDC estimates. Similarly, for 2021 and 2022, we projected that there would be 105,450 and 117,430 PWH 65y+ in care, compared to 100,440 and 112,574 respectively **(eTable 10)**.

*Projected deaths (2026-2035)*

We projected that the total number of deaths of Medicare beneficiaries with HIV on ART would be 5,500 in 2026 and 9,640 in 2035. Increased number of projected deaths in future years is commensurate with the increased population sizes: 1,600 to 1,950 (65-69y), 1,450 to 2,520 (70-74y), 1,060 to 2,420 (75-79y), and 1,400 to 2,760 (80y+) in 2026 and 2035, respectively **(eTable 11).**

*Impact of cost parameterization*

We parameterized costs in four distinct ways: 1) adjusting for higher costs among Medicare Advantage beneficiaries compared with Traditional Medicare beneficiaries, as well as inflation (base case); 2) adjusting for higher costs among Medicare Advantage beneficiaries but not for inflation; 3) adjusting for inflation but not higher costs among Medicare Advantage beneficiaries; and 4) adjusting for neither **(eTable 12).**

*One-way sensitivity analysis of ART costs*

Given the impact of annual ART costs on total Medicare spending for PWH, we performed a one-way sensitivity analysis on ART costs to project 10-year cumulative costs **(eFigure 1)**. If ART costs are reduced to 60% of 2023 ART costs, cumulative costs over the next 10 years would be $117.5B, which is 39.9% lower than base case projections of cumulative costs. If ART costs increase by 20%, then 10-year cumulative costs would reach $221.6B by 2035.

*Additional sensitivity analyses: population outcomes*

We performed a multiway sensitivity analysis to assess the impact of varying non-HIV-related mortality and the number of Medicare beneficiaries 65y+ on ART in 2026 on the projected population size of Medicare beneficiaries aged 65y+ on ART in 2035. When non-HIV-related mortality is 20% higher than in the base case, and the number of Medicare beneficiaries 65y+

on ART in 2026 is 10% lower than in the base case, the projected population size is 178,290. When non-HIV-related mortality is 20% lower than in base case and the number of Medicare beneficiaries 65y+ on ART in 2026 is 10% higher than in base case, the projected population size is 211,240 **(eFigure 2).**

*Additional sensitivity analyses: cost outcomes*

We performed a range of multiway sensitivity analyses to assess the impact on 10-year cumulative costs for Medicare beneficiaries with HIV who are 65y+. When the percent of PWH eligible for Medicare is 94% and background mortality is 20% lower than in base case, the projected cumulative costs are $255.7B, as compared to $174.8B if the percent of PWH eligible is 68% and background mortality is 20% higher than in the base case **(eFigure 3A).** Estimated ART costs are particularly influential in 10-year cumulative costs, even when also varying the the number of Medicare-eligible PWH who are turning 65 years old annually (**eFigure 3B**) or the number of Medicare beneficiaries who are 65y+ in 2026 **(eFigure 3C).** Comparatively, non-HIV-related costs are less influential, even when varying the percent of PWH 65y+ in care who are enrolled in Medicare **(eFigure 3D).** When the cost of ART is 40% of 2023 costs and the number of Medicare-eligible PWH turning age 65 is 90% of the base case value; the 10-year cumulative costs is $112.1B, as compared to $232.0B when the cost of ART is 20% higher than 2023 costs and the number of Medicare-eligible PWH turning 65 is 10% higher than in base case. A similar pattern is observed when varying the cost of ART and the number of Medicare beneficiaries who are 65y+ in 2026; at 40% of 2023 ART costs and 90% of Medicare beneficiaries who are 65y+ in 2026, total costs are projected to be $185.5B, as compared to at 120% of 2023 ART costs and 110% of Medicare beneficiaries 65y+, where total costs are $205.6B. Finally, when non-HIV-related costs are 98% of 2023 costs, and the percent of PWH 65y+ in care who are enrolled in Medicare is 68%, model-projected costs are $177.3 billion, as compared to $251.7

billion at 102% of 2023 non-HIV-related costs and 94% of Medicare beneficiaries 65y+ in care being enrolled in Medicare.

38. Walmsley S, Baumgarten A, Berenguer J, et al. Brief report: dolutegravir plus abacavir/lamivudine for the treatment of HIV-1 infection in antiretroviral therapy-naive

patients: week 96 and week 144 results from the single randomized clinical trial. *J Acquir Immune Defic Syndr*. 2015;70(5):515-519. doi:10.1097/QAI.0000000000000790

39. Raffi F, Rachlis A, Stellbrink HJ, et al. Once-daily dolutegravir versus raltegravir in antiretroviral-naive adults with HIV-1 infection: 48 week results from the randomised, double-blind, non-inferiority SPRING-2 study. *The Lancet*. 2013;381(9868):735-743. doi:10.1016/S0140-6736(12)61853-4

40. Clotet B, Feinberg J, van Lunzen J, et al. Once-daily dolutegravir versus darunavir plus ritonavir in antiretroviral-naive adults with HIV-1 infection (FLAMINGO): 48 week results from the randomised open-label phase 3b study. *The Lancet*. 2014;383(9936):2222-2231. doi:10.1016/S0140-6736(14)60084-2

41. Sax PE, Pozniak A, Montes ML, et al. Coformulated bictegravir, emtricitabine, and tenofovir alafenamide versus dolutegravir with emtricitabine and tenofovir alafenamide, for initial treatment of HIV-1 infection (GS-US-380–1490): a randomised, double-blind, multicentre, phase 3, non-inferiority trial. *The Lancet*. 2017;390(10107):2073-2082. doi:10.1016/S0140-6736(17)32340-1

42. Orrell C, Hagins DP, Belonosova E, et al. Fixed-dose combination dolutegravir, abacavir, and lamivudine versus ritonavir-boosted atazanavir plus tenofovir disoproxil fumarate and emtricitabine in previously untreated women with HIV-1 infection (ARIA): week 48 results from a randomised, open-label, non-inferiority, phase 3b study. *Lancet HIV*. 2017;4(12):e536-e546. doi:10.1016/S2352-3018(17)30095-4

43. Gallant J, Lazzarin A, Mills A, et al. Bictegravir, emtricitabine, and tenofovir alafenamide versus dolutegravir, abacavir, and lamivudine for initial treatment of HIV-1 infection (GS-US-

380-1489): a double-blind, multicentre, phase 3, randomised controlled non-inferiority trial. *The Lancet*. 2017;390(10107):2063-2072. doi:10.1016/S0140-6736(17)32299-7

49. Centers for Medicare and Medicaid Services. Chronic Conditions Data Warehouse. https://www2.ccwdata.org/web/guest/medicare-data

50. HHS ASPE. Analysis of new generic markets, Effect of market entry on generic drug prices: Medicare data 2007-2022. https://aspe.hhs.gov/sites/default/files/documents/510e964dc7b7f00763a7f8a1dbc5ae7b/aspe-ib-generic-drugs-competition.pdf

51. Centers for Medicare and Medicaid Services. Medicare drug price negotiation program: Initial memorandum, implementation of sections 1191-1198 of the Social Security Act for initial price applicability year 2026, and solicitation of comments. Published online March 15, 2023. Accessed December 15, 2025. https://www.cms.gov/files/document/medicare-drug-price-negotiation-program-initial-guidance.pdf

**eTable 1.** CHEERS 2022 Checklist

| Topic | No. | Item | Location where item is reported |
|---|---|---|---|
| **Title** | 1 | Identify the study as an economic evaluation and specify the interventions being compared. | Title (Page 1) |
| **Abstract** | 2 | Provide a structured summary that highlights context, key methods, results, and alternative analyses. | Abstract (Page 4) |
| **Introduction** | | | |
| **Background and objectives** | 3 | Give the context for the study, the study question, and its practical relevance for decision making in policy or practice. | Introduction (Page 6) |
| **Methods** | | | |
| **Health economic analysis plan** | 4 | Indicate whether a health economic analysis plan was developed and where available. | Not applicable |
| **Study population** | 5 | Describe characteristics of the study population (such as age range, demographics, socioeconomic, or clinical characteristics). | Methods (Pages 7-9) |
| **Setting and location** | 6 | Provide relevant contextual information that may influence findings. | Introduction (Page 6) |
| **Comparators** | 7 | Describe the interventions or strategies being compared and why chosen. | Not applicable |

| Topic | No. | Item | Location where item is reported |
| --- | --- | --- | --- |
| **Perspective** | 8 | State the perspective(s) adopted by the study and why chosen. | Introduction; Page 6 |
| **Time horizon** | 9 | State the time horizon for the study and why appropriate. | Methods (page 7) |
| **Discount rate** | 10 | Report the discount rate(s) and reason chosen. | Methods (page 10) |
| **Selection of outcomes** | 11 | Describe what outcomes were used as the measure(s) of benefit(s) and harm(s). | Methods (page 7) |
| **Measurement of outcomes** | 12 | Describe how outcomes used to capture benefit(s) and harm(s) were measured. | Methods (page 7) |
| **Valuation of outcomes** | 13 | Describe the population and methods used to measure and value outcomes. | Methods (page 7) |
| **Measurement and valuation of resources and costs** | 14 | Describe how costs were valued. | Methods (pages 9-10); eMethods |
| **Currency, price date, and conversion** | 15 | Report the dates of the estimated resource quantities and unit costs, plus the currency and year of conversion. | Methods (Pages 9-10) |
| **Rationale and description of model** | 16 | If modelling is used, describe in detail and why used. Report if the model is publicly available and where it can be accessed. | Introduction; Page 6 |

| Topic | No. | Item | Location where item is reported |
|---|---|---|---|
| **Analytics and assumptions** | 17 | Describe any methods for analysing or statistically transforming data, any extrapolation methods, and approaches for validating any model used. | Methods (Page 7-10) eMethods, eTables 10-12 |
| **Characterising heterogeneity** | 18 | Describe any methods used for estimating how the results of the study vary for subgroups. | Not applicable |
| **Characterising distributional effects** | 19 | Describe how impacts are distributed across different individuals or adjustments made to reflect priority populations. | Not applicable |
| **Characterising uncertainty** | 20 | Describe methods to characterise any sources of uncertainty in the analysis. | Methods (Page 10) and eTable 7 |
| **Approach to engagement with patients and others affected by the study** | 21 | Describe any approaches to engage patients or service recipients, the general public, communities, or stakeholders (such as clinicians or payers) in the design of the study. | Not applicable |
| **Results** | | | |
| **Study parameters** | 22 | Report all analytic inputs (such as values, ranges, references) including uncertainty or distributional assumptions. | Methods; Page 7-9; eTables 1-7 |

| Topic | No. | Item | Location where item is reported |
|---|---|---|---|
| **Summary of main results** | 23 | Report the mean values for the main categories of costs and outcomes of interest and summarise them in the most appropriate overall measure. | Key points (Page 3), Abstract (Page 4) and Table 2 |
| **Effect of uncertainty** | 24 | Describe how uncertainty about analytic judgments, inputs, or projections affect findings. Report the effect of choice of discount rate and time horizon, if applicable. | Results; Pages 11-13 |
| **Effect of engagement with patients and others affected by the study** | 25 | Report on any difference patient/service recipient, general public, community, or stakeholder involvement made to the approach or findings of the study | Not applicable |
| **Discussion** | | | |
| **Study findings, limitations, generalizability, and current knowledge** | 26 | Report key findings, limitations, ethical or equity considerations not captured, and how these could affect patients, policy, or practice. | Discussion; Pages 15-16 |
| **Other relevant information** | | | |
| **Source of funding** | 27 | Describe how the study was funded and any role of the funder in the identification, design, conduct, and reporting of the analysis | Page 2 |

| Topic | No. | Item | Location where item is reported |
|---|---|---|---|
| **Conflicts of interest** | 28 | Report authors conflicts of interest according to journal or International Committee of Medical Journal Editors requirements. | Page 2 |

*From:* Husereau D, Drummond M, Augustovski F, et al. Consolidated Health Economic Evaluation Reporting Standards 2022 (CHEERS 2022) Explanation and Elaboration: A Report of the ISPOR CHEERS II Good Practices Task Force. Value Health 2022;25. doi:10.1016/j.jval.2021.10.008

**eTable 2.** Input parameters for the simulated cohorts of PWH age 20y+ in the United States in the CEPAC-US Model.

| Parameter | Base Case Input | Reference |
|---|---|---|
| Baseline cohort characteristics | | |
| Male (%) | 77 | [4] |
| Initial CD4 count, mean cells/µl (SD) | | |
| Prevalent, undiagnosed | 400 (166) | [30] |
| Prevalent, on ART | 576 (227) | [31] |
| Prevalent, disengaged from care | 325 (53) | [30] |
| Incident | 652 (101) | [32] |
| HIV RNA **setpoint** distribution (%) | | [33] |
| >100,000 copies/ml | 25 | |
| 30,001-100,000 copies/ml | 42 | |
| 10,001-30,000 copies/ml | 21 | |
| 3,001-10,000 copies/ml | 6 | |
| ≤ 3,000 copies/ml | 6 | |
| Initial age among people with prevalent HIV at model start in Jan 2015 (%) | | [6,13,34,35] |
| 20-24 years | 6 | |
| 25-34 years | 17 | |
| 35-44 years | 21 | |
| 45-54 years | 31 | |
| 55-64 years | 19 | |
| 65-69 years | 3 | |
| ≥ 70 years | 2 | |

**eTable 2.** Input parameters for the simulated cohorts of PWH age 20y+ in the United States in the CEPAC-US Model. (continued)

| Parameter | Base Case Input | Reference |
|---|---|---|
| **Baseline cohort characteristics** | | |
| Initial age among people newly infected with HIV (%) | *From 2015 onwards* | [7,34] |
| 20-24 years | 24.4 | |
| 25-34 years | 35.8 | |
| 35-44 years | 18.3 | |
| 45-54 years | 14.2 | |
| 55-64 years | 5.8 | |
| 65-69 years | 0.8 | |
| ≥ 70 years | 0.6 | |
| Population size, n | | [6,7,34] |
| Prevalent, undiagnosed | 151,954 | |
| Prevalent, on ART | 675,439 | |
| Prevalent, disengaged from care | 244,778 | |
| Incident | Determined by transmissions | |
| **HIV Testing** | | |
| Monthly HIV testing probability (%) | | |
| Prevalent, undiagnosed | 1.2 | Assumption |
| Incident | 1.7 | Assumption |

**eTable 2.** Input parameters for the simulated cohorts of PWH age 20y+ in the United States in the CEPAC-US Model. (continued)

| Parameter | Base Case Input | Reference |
|---|---|---|
| Monthly mortality probabilities | | |
| Associated with OIs | | 36 |
| *Pneumocystis* pneumonia (PCP) | 0.035 | |
| *Mycobacterium avium* complex (MAC) | 0.045 | |
| Toxoplasmosis | 0.182 | |
| Cytomegalovirus (CMV) | 0.048 | |
| Fungal infection | 0.036 | |
| Other OI | 0.043 | |
| Associated with chronic HIV, stratified by CD4 count | | 36 |
| ≥ 500 cells/µl | 0.000033 | |
| 350-500 cells/µl | 0.000105 | |
| 200-350 cells/µl | 0.000212 | |
| 100-200 cells/µl | 0.000204 | |
| 50-100 cells/µl | 0.000281 | |
| ≤ 50 cells/µl | 0.001207 | |
| Associated with chronic HIV, stratified by CD4 count | | |
| ≥ 500 cells/µl | 0.000033 | |
| 350-500 cells/µl | 0.000105 | |
| 200-350 cells/µl | 0.000212 | |
| 100-200 cells/µl | 0.002700 | |
| 50-100 cells/µl | 0.002660 | |
| ≤ 50 cells/µl | 0.005644 | |

**eTable 2.** Input parameters for the simulated cohorts of PWH age 20+ in the United States in the CEPAC-US Model. (continued)

| Parameter | Base Case Input | Reference |
|---|---|---|
| **Monthly Mortality Probabilities** | | |
| Non-HIV-related causes of death | | [37] |
| Male at birth | | |
| 20-24 years | 0.000162 | |
| 25-34 years | 0.000211 | |
| 35-44 years | 0.000268 | |
| 45-54 years | 0.000484 | |
| 55-64 years | 0.000954 | |
| 65-69 years | 0.001900 | |
| ≥ 70 years | 0.002760 | |
| Female at birth | | |
| 20-24 years | 0.000152 | |
| 25-34 years | 0.000210 | |
| 35-44 years | 0.000371 | |
| 45-54 years | 0.000799 | |
| 55-64 years | 0.000951 | |
| 65-69 years | 0.001942 | |
| ≥ 70 years | 0.003029 | |
| **ART efficacy** | | |
| % Suppressed at 12 months if 95% adherence | | |
| INSTI-based regimen | 96.4 | Derived from [38–42] |
| Monthly probability of late Failure (%) | 0.20 | Derived from [38–40,43] |

**eTable 2.** Input parameters for the simulated cohorts of PWH age 20y+ in the United States in the CEPAC-US Model. (continued)

| Parameter | Base Case Input | Reference |
| --- | --- | --- |
| HIV transmission rate by disease stage and viral load, transmissions/100 person years | | [44,45] |
| Acute infection (3 months after infection) | 65.47 | |
| Late-stage disease (CD4 cell count <200 cells/μl) | 9.03 | |
| > 100,000 copies/mL | 9.03 | |
| 10,001 – 100,000 copies/mL | 8.12 | [44,45] |
| 3,001 – 10,000 copies/mL | 4.17 | |
| 501 – 3,000 copies/mL | 2.06 | |
| 20 – 500 copies/mL | 0.16 | |
| ≤ 20 copies/mL | 0 | |
| Engagement in care | | |
| Monthly probability of disengagement from care (%) | 0.01-15 | Derived from [46,47] |
| Monthly probability of return to care (%) | 3.0 | Derived from [48] |
| Propensity to respond coefficients* | Age-adjusted | [2] |

*As previously described, we developed age-stratified likelihood of virologic suppression and engagement in care.[2]

**eTable 3.** Calibration of model transmissions to CDC surveillance estimates.

| Year | CDC Estimated HIV Incidence [a] [29] | CDC 95% Confidence Interval [29] | HIV Transmission Deflator | Model-Projected HIV Incidence |
|---|---|---|---|---|
| 2015 | 37,100 | 36,000 - 38,200 | 1 | 36,590 |
| 2016 | 37,200 | 36,000 - 38,400 | 0.95 | 38,160 |
| 2017 | 36,200 | 34,800 - 37,500 | 0.90 | 37,660 |
| 2018 | 36,200 | 34,600 - 37,700 | 0.85 | 36,100 |
| 2019 | 35,100 | 33,400 - 36,800 | 0.80 | 34,050 |
| 2020 | 34,200 | 32,100 - 36,300 | 0.80 | 34,190 |
| 2021 | 32,700 | 30,400 - 34,900 | 0.75 | 31,940 |
| 2022 | 31,800 | 29,200 - 34,400 | 0.75 | 31,870 |

[a] CDC estimates are reported for PWH 13y+, whereas the CEPAC model projects PWH 20y+.

**eTable 4.** Mean annual per-person costs by category while alive and in the year of death for male Medicare beneficiaries with HIV on ART for 10-12 months of the year.[49]

| | ART ($) | | | HIV-related care (excluding ART) ($) | | | Non-HIV-related care ($) | | |
|---|---|---|---|---|---|---|---|---|---|
| **Mean annual per-person costs in each year survived ($)** | | | | | | | | | |
| Age (years) | TM | MA | TM+MA | TM | MA | TM+MA | TM | MA | TM+MA |
| 65-69 | 46,337 | 56,531 | 52,405 | 1,706 | 2,081 | 1,930 | 24,012 | 29,295 | 27,156 |
| 70-74 | 44,938 | 54,824 | 50,823 | 1,610 | 1,964 | 1,821 | 23,864 | 29,114 | 26,989 |
| 75-79 | 43,465 | 53,027 | 49,157 | 1,862 | 2,272 | 2,106 | 26,016 | 31,740 | 29,423 |
| ≥ 80 | 42,440 | 51,777 | 47,998 | 2,523 | 3,078 | 2,854 | 28,772 | 35,102 | 32,540 |
| **Mean per-person costs in 12 months prior to death ($)** | | | | | | | | | |
| 65-69 | 46,002 | 56,122 | 51,870 | 14,648 | 17,871 | 16,516 | 86,251 | 105,226 | 97,252 |
| 70-74 | 44,400 | 54,168 | 50,063 | 15,524 | 18,939 | 17,504 | 81,770 | 99,759 | 92,200 |
| 75-79 | 40,925 | 49,929 | 46,145 | 13,446 | 16,404 | 15,161 | 89,176 | 108,795 | 100,550 |
| ≥ 80 | 41,394 | 50,501 | 46,674 | 11,987 | 14,624 | 13,516 | 71,213 | 86,880 | 80,296 |

Abbreviations: TM: Traditional Medicare beneficiaries, MA: Medicare Advantage beneficiaries, TM+MA: Traditional Medicare and Medicare Advantage beneficiaries overall

**eTable 5.** Mean annual per-person costs by category while alive and in the year of death for female Medicare beneficiaries with HIV on ART for 10-12 months of the year.[49]

| | ART ($) | | | HIV-related care (excluding ART) ($) | | | Non-HIV-related care ($) | | |
|---|---|---|---|---|---|---|---|---|---|
| **Mean annual per-person costs in year survived ($)** | | | | | | | | | |
| Age (years) | TM | MA | TM+MA | TM | MA | TM+MA | TM | MA | TM+MA |
| 65-69 | 46,747 | 57,031 | 53,668 | 1,668 | 2,035 | 1,915 | 22,569 | 27,534 | 25,911 |
| 70-74 | 44,245 | 53,979 | 50,796 | 1,753 | 2,138 | 2,012 | 22,339 | 27,254 | 25,647 |
| 75-79 | 43,970 | 53,643 | 50,480 | 2,060 | 2,513 | 2,365 | 22,587 | 27,556 | 25,931 |
| ≥ 80 | 42,808 | 52,226 | 49,146 | 2,137 | 2,607 | 2,453 | 23,936 | 29,202 | 27,480 |
| **Mean per-person costs in 12 months prior to death ($)** | | | | | | | | | |
| 65-69 | 42,611 | 51,985 | 48,514 | 18,514 | 22,587 | 21,079 | 88,300 | 107,726 | 100,533 |
| 70-74 | 42,270 | 51,569 | 48,126 | 11,190 | 13,652 | 12,740 | 77,761 | 94,868 | 88,534 |
| 75-79 | 41,567 | 50,712 | 47,326 | 13,534 | 16,511 | 15,409 | 61,138 | 74,588 | 69,608 |
| ≥ 80 | 41,727 | 50,907 | 47,508 | 15,219 | 18,567 | 17,327 | 80,343 | 98,018 | 91,473 |

Abbreviations: TM: Traditional Medicare beneficiaries, MA: Medicare Advantage beneficiaries, TM+MA: Traditional Medicare and Medicare Advantage beneficiaries overall.

**eTable 6.** Personal health care index projections used to adjust HIV-related non-ART and non-HIV-related care costs.

| Year | CMS personal healthcare price indices [26] | Ratio (Base = 2023) |
|---|---|---|
| 2023 | 112.7 | 1 |
| 2024 | 115.9 | 1.03 |
| 2025 | 119.0 | 1.06 |
| 2026 | 122.3 | 1.09 |
| 2027 | 125.8 | 1.12 |
| 2028 | 129.3 | 1.15 |
| 2029 | 132.9 | 1.18 |
| 2030 | 136.5 | 1.21 |
| 2031 | 140.1 | 1.24 |
| 2032 | 143.9 | 1.28 |
| 2033 | 147.6 | 1.31 |
| 2034* | 150.7 | 1.34 |
| 2035* | 154.3 | 1.37 |

*The personal healthcare index for 2024-2033 are projected by the Centers for Medicare and Medicaid Services. For 2034 and 2035, we projected the CPI based on the trend in projected price indices between 2024-2033.

**eTable 7.** Parameters assessed in sensitivity analysis: Base case values, lower-bound (LB), upper-bound (UB) and justifications for ranges

| Input | Base case | LB | Reference | UB | Reference |
|---|---|---|---|---|---|
| **Population parameters** | | | | | |
| Percent of PWH enrolled in Medicare | 74 | 68 | Derived[a] from Ryan White[22] | 94 | Derived from Census.gov data for general 65y+ pop[23] |
| Number of PWH in care turning 65 | 694-925 | 90% | Assumption | 110% | Assumption |
| Number of PWH in care at model start in Jan 2026 | 150,780 | 90% | Assumption | 110% | Assumption |
| Number of PWH age 65y+ newly diagnosed and entering care monthly | 37 | 90% | Assumption | 110% | Assumption |
| Non-HIV-related mortality | Age-/sex-stratified | 80% | Assumption | 120% | Assumption |
| **Costs parameters** | | | | | |
| ART costs | $48,296-$52,733 | 40% | Estimated impact of generic entry; Maximum minimum discount proposed by IRA[50,51] | 120% | Assumption |
| Medical costs in the year of death | See eTable 8 | 90% | 95% CI from CMS cost data | 110% | Assumption |
| Non-HIV-related costs | $26,833-31,226 | 98% | 95% CI from CMS cost data | 102% | 95% CI from CMS cost data |
| Non-ART HIV-related costs | $1,871-2,750 | 95% | 95% CI from CMS cost data | 105% | 95% CI from CMS cost data |

[a]See technical appendix section *Projecting the number of PWH age 65y+ enrolled in Medicare*

**eTable 8.** Model-projected numbers of deaths among Medicare beneficiaries with HIV aged 65y+ by year-end.

| **Number of deaths among Medicare beneficiaries with HIV aged 65y+ by year-end** | | | | | |
|---|---|---|---|---|---|
| **Year** | **65-69y** | **70-74y** | **75-79y** | **80y+** | **All 65y+** |
| 2026 | 1,600 | 1,450 | 1,060 | 1,400 | 5,500 |
| 2027 | 1,680 | 1,540 | 1,180 | 1,450 | 5,850 |
| 2028 | 1,740 | 1,670 | 1,290 | 1,470 | 6,180 |
| 2029 | 1,820 | 1,810 | 1,480 | 1,540 | 6,650 |
| 2030 | 1,910 | 1,960 | 1,610 | 1,590 | 7,070 |
| 2031 | 1,970 | 2,110 | 1,740 | 1,770 | 7,590 |
| 2032 | 1,980 | 2,180 | 1,910 | 2,030 | 8,100 |
| 2033 | 1,980 | 2,260 | 2,060 | 2,300 | 8,600 |
| 2034 | 1,970 | 2,380 | 2,230 | 2,550 | 9,140 |
| 2035 | 1,950 | 2,520 | 2,420 | 2,760 | 9,640 |

**eTable 9.** Comparison of model-projected costs (in billions of US dollars) based on 4 different approaches to adjusting for higher costs among Medicare Advantage beneficiaries (vs Traditional Medicare beneficiaries) and/or for inflation (vs not).

| Year | MA + Inflation Adjustment (base case) | MA Adjustment only | Inflation adjustment only | No MA or inflation adjustment |
|---|---|---|---|---|
| 2026 | $11.4B | $9.9B | $10.0B | $8.7B |
| 2027 | $13.0B | $10.8B | $11.4B | $9.5B |
| 2028 | $14.6B | $11.6B | $12.9B | $10.2B |
| 2029 | $16.4B | $12.4B | $14.5B | $10.9B |
| 2030 | $18.3B | $13.1B | $16.1B | $11.6B |
| 2031 | $20.2B | $13.9B | $17.8B | $12.2B |
| 2032 | $22.2B | $14.6B | $19.6B | $12.9B |
| 2033 | $24.4B | $15.2B | $21.5B | $13.4B |
| 2034 | $26.5B | $15.8B | $23.4B | $14.0B |
| 2035 | $28.6B | $16.3B | $25.2B | $14.4B |
| Total | $195.6B | $133.6B | $172.4B | $117.8B |

Abbreviation: B: billion

**eTable 10**. Validation of CEPAC model output of PWH living with HIV and deaths among PWH diagnosed with HIV aged 20y+ from 2017-2022.

| | Persons living with diagnosed HIV infection age 20y or older | | | Deaths of persons with diagnosed HIV infection age 20y or older | | |
|---|---|---|---|---|---|---|
| Year | CDC Data | Model output | Reference | CDC Data | Model output | Reference |
| 2017 | 988,605 | 992,700 | [4] | 16,336 | 14,700 | [12] |
| 2018 | 1,009,449 | 1,016,450 | | 16,183 | 16,300 | |
| 2019 | 1,031,893 | 1,037,700 | | 16,254 | 17,740 | |
| 2020 | 1,045,249 | 1,056,380 | [11] | 19,123 | 18,960 | [11] |
| 2021 | 1,063,805 | 1,072,660 | | 20,176 | 19,970 | |
| 2022 | 1,087,872 | 1,086,840 | | 18,931 | 20,730 | |

**eTable 11.** Validation of CEPAC model output of receipt of care among diagnosed PWH 20y+ and virologic suppression among PWH 20y+ in care in 2022.

| | CDC Estimates | Model output | Reference |
|---|---|---|---|
| Receipt of HIV medical care among PWH diagnosed by year-end 2021 and alive at year-end 2022 (%) [a] | 70.7 | 70.6 | [10] |
| Virologically suppressed among PWH diagnosed by year-end 2021 and alive at year-end 2022 (%) | 63.7 | 67.3 | [10] |

[a] For CDC estimates, we defined receipt of HIV medical care as at least one CD4 count or VL test during 2022.

**eTable 12.** Validation of CHARMED model output for the number of PWH over the age of 65, diagnosed, and in care compared with CDC data (2021-2023).

| Year | CDC PWH 65y+ living with diagnosed HIV[29] | CDC (% of diagnosed in care)[29] | Estimated number of PWH 65y+ in care[a] | Model projections |
|---|---|---|---|---|
| 2021 | 142,670 | 70.4 | 100,440 | 105,450 |
| 2022 | 159,227 | 70.7 | 112,574 | 117,430 |
| 2023[b] | 176,647 | 71.0 | 125,419 | 128,780 |

[a] CDC estimates for the number of people diagnosed 65y+ and percent in care are not directly comparable. While the estimate for diagnosed prevalence includes all PWH alive at the end of the specified year, the estimate for percent in care includes only people who were diagnosed by the end of the previous year. To address this, we derived the estimated number of PWH in care by taking the estimated percent in care for a given year and applying it to the total population of PWH 65y+ diagnosed.

[b] CDC data for 2023 are considered preliminary.

### FIGURE LEGENDS

**eFigure 1. Impact of changes in ART costs on 10 years of Medicare spending for beneficiaries aged 65y+ on ART (2026-2035).**

The x-axis displays the percentage of the 2023 ART costs, ranging from 40% (a 60% reduction, left) to 120% (a 20% increase, right). Light blue represents other non-HIV-related costs, dark blue represents costs related to direct medical treatment of HIV, and grey represents ART costs. Abbreviations: ART: antiretroviral therapy.

**eFigure 2. Multi-way sensitivity analysis of non-HIV-related mortality and the number of Medicare beneficiaries on ART in 2026.**

The model-projected number of Medicare beneficiaries with HIV who are on ART and 65y+ in 2035 are displayed given a range in non-HIV-related mortality rates with the number of Medicare beneficiaries 65y+ on ART in 2026. Base case results are designated with an "X." Light orange shows lower 10-year population size, whereas dark blue shows higher 10-year population size. Abbreviations: ART: antiretroviral therapy.

**eFigure 3. Additional multi-way sensitivity analyses comparing the total cumulative projected costs of Medicare beneficiaries aged 65y+ on ART between 2026 and 2035.**

The model-projected 10-year cumulative costs are displayed given a range of non-HIV-related mortality and the percent of PWH 65y+ eligible for Medicare (Panel A), percent of 2023 ART costs and the number of Medicare beneficiaries with HIV 65 years and older on ART in 2026 (Panel B), and percent of 2023 ART costs and the number of Medicare-eligible PWH in care turning age 65 (Panel C). Base case results are designated with an "X." Light blue shows lower 10-year costs, whereas dark blue shows higher 10-year costs. Abbreviations: PWH: people with HIV; ART: antiretroviral therapy.

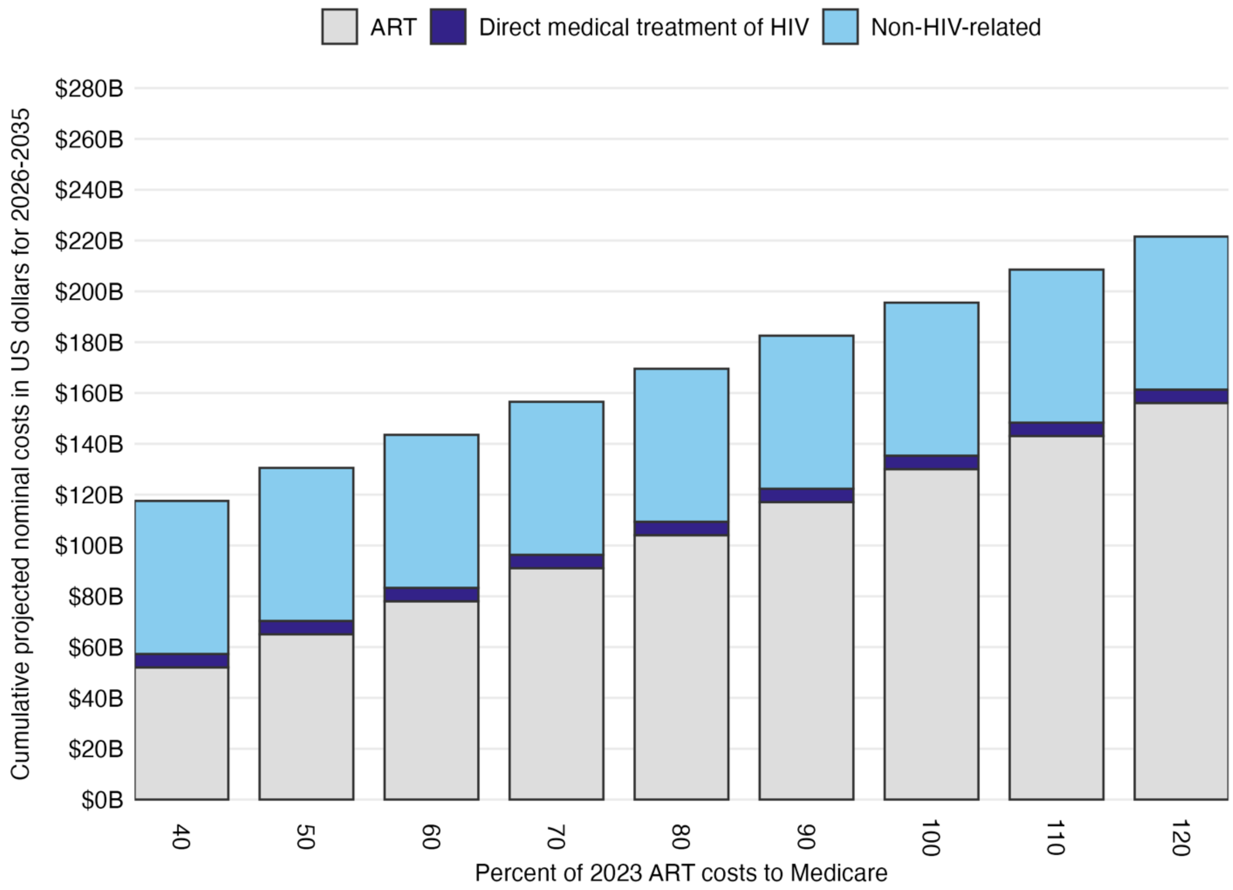


**eFigure 1.** Impact of changes in ART costs on 10 years of Medicare spending for beneficiaries aged 65y+ on ART (2026-2035).

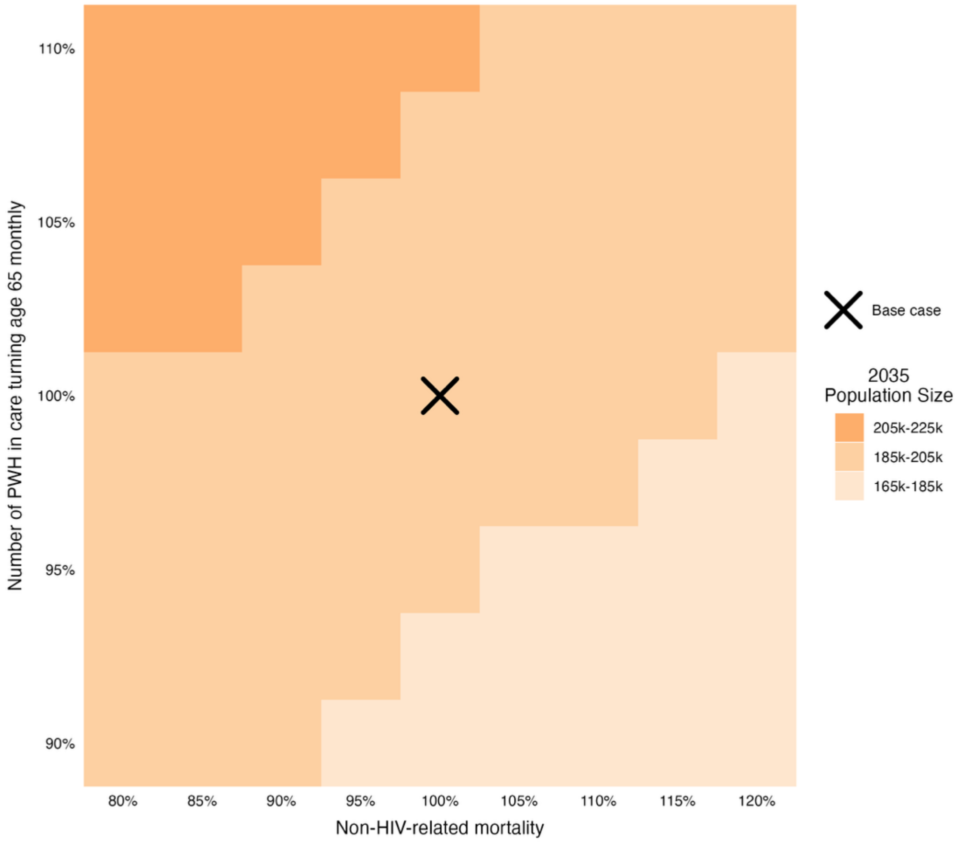

**eFigure 2.** Multi-way sensitivity analysis of non-HIV-related mortality and the number of Medicare beneficiaries on ART in 2026.

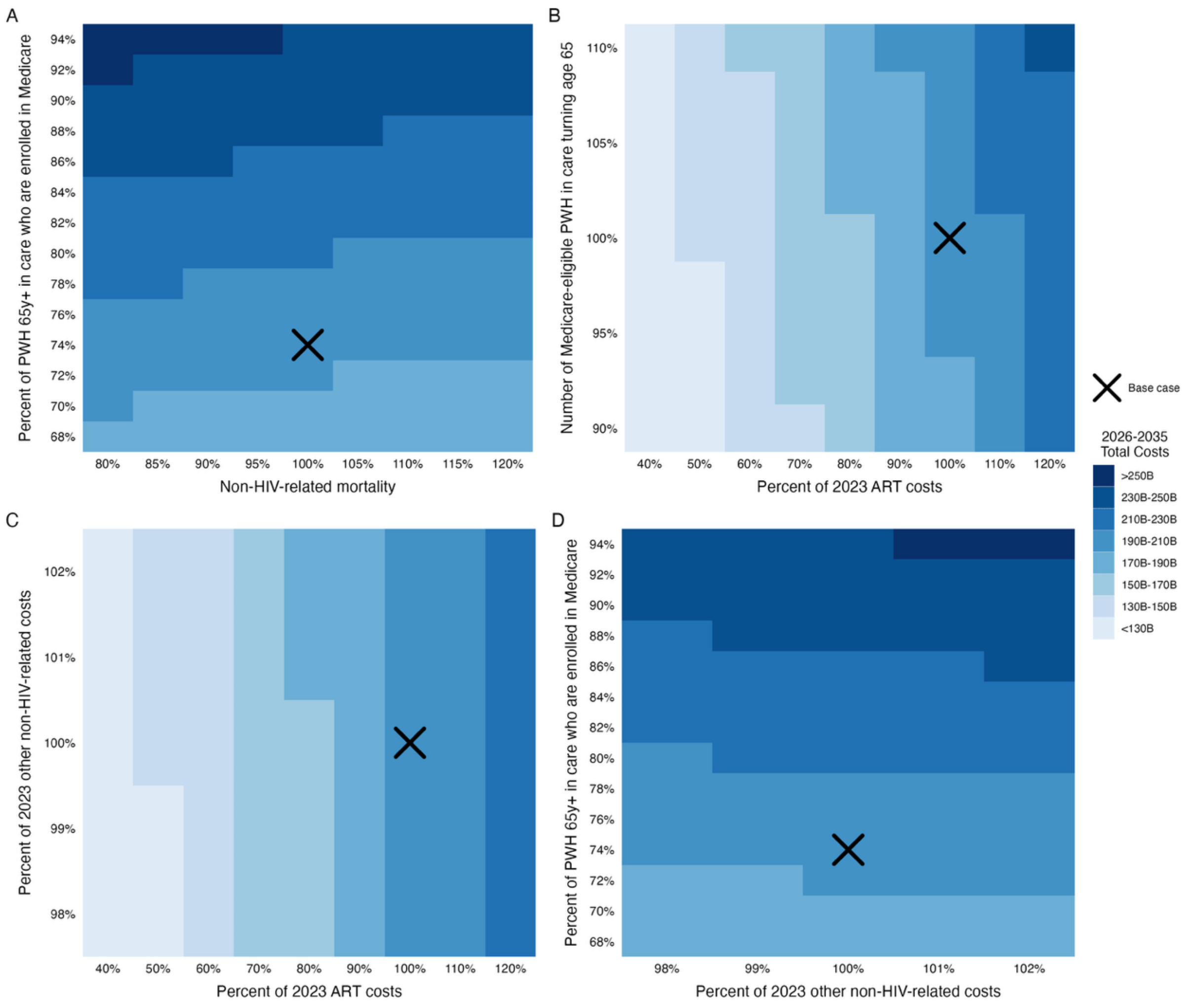


**eFigure 3.** Additional multi-way sensitivity analyses comparing the total cumulative projected costs of Medicare beneficiaries aged 65y+ on ART between 2026 and 2035.
